# Machine Learning Approaches for Survival Prediction in Bladder Cancer: A Single-Center Analysis of Clinical and Inflammatory Markers

**DOI:** 10.1101/2024.11.26.24317989

**Authors:** Francesco Andrea Causio, Vittorio De Vita, Andrea Nappi, Melissa Sawaya, Bernardo Rocco, Nazario Foschi, Giuseppe Maioriello, Pierluigi Russo

## Abstract

This study investigated the application of machine learning algorithms for survival prediction in bladder cancer patients undergoing cystectomy. We analyzed retrospective data from 370 patients, developing predictive models for disease-free survival (DFS), overall survival (OS), and cause of death. Multiple machine learning approaches were employed, including linear regression, random forests, gradient boosting, support vector machines, and neural networks. The models achieved mean absolute errors of 22-23 months for survival predictions and 66.67% accuracy in cause-of-death classification. Clinical T-stage emerged as the strongest predictor, while the Systemic Immune-Inflammation Index (SII) demonstrated a consistent negative correlation with survival outcomes. An unexpected positive correlation between age and survival was observed, possibly reflecting selection bias in surgical candidates. The study’s findings suggest that machine learning approaches, despite current limitations, offer promising tools for risk stratification in clinical trial design and patient allocation, though further refinement is needed for individual prognostication.

## Introduction

In the evolving landscape of healthcare, the integration of artificial intelligence (AI) into clinical decision-making has gained significant momentum, particularly in the realm of oncology. (1–3) With advancements in machine learning techniques, healthcare professionals are increasingly harnessing the power of AI to enhance diagnosis, prognosis, and treatment planning. This rise in predictive algorithms is fueled by the exponential growth of digital healthcare data, including electronic health records, medical imaging, genomic data, and real-time patient monitoring. (3,4)

The field of urology is complex: cancerous conditions benefit from the leverage of additional data sources and decision-making algorithms that allow physicians to plan treatment while considering several complex factors. Urological cancers, including prostate, bladder, and renal cancers, pose a considerable burden on healthcare systems globally. (5) These malignancies often require complex management involving early diagnosis, accurate staging, and personalized treatment strategies to optimize patient outcomes. Traditional methods of assessing prognosis rely heavily on statistical models that may not capture the multifaceted nature of cancer behavior and patient responses to treatment. As highlighted in the review by Abbod et al., conventional regression statistics often fall short of providing the depth of analysis required to address the complexities of cancer management. In contrast, AI techniques, such as artificial neural networks (ANNs), Bayesian networks, and neuro-fuzzy modeling systems, offer innovative approaches to constructing data-driven models that can adapt to the heterogeneous nature of cancer. (6)

The potential of AI in predicting patient outcomes is particularly evident in its ability to analyze large datasets without the constraints of predetermined statistical distributions. By leveraging retrospective data, we can develop algorithms that not only identify patterns and correlations but also provide insights into individual patient behavior. This capability is crucial for clinicians who face the challenge of tailoring treatment plans to the unique characteristics of each patient. In the context of mortality and post-operative survival, the application of AI can provide critical insights that enhance our understanding of patient outcomes following surgical interventions. The ability to predict which patients are at higher risk of complications or recurrence can lead to more informed clinical decisions, ultimately improving the quality of care. (7) For instance, machine learning algorithms can analyze a multitude of variables, including clinical, pathological, and demographic factors, to generate individualized risk profiles that guide treatment strategies and follow-up plans. (8)

In this study, we will focus specifically on the training of an AI algorithm using retrospective data collected from patients diagnosed with bladder cancer and undergoing cystectomy. Patients with localized muscle-invasive or recurrent non-muscle invasive bladder cancer benefit most from radical cystectomy preceded in some patients by neoadjuvant chemotherapy in terms of local control. Even with sufficient local control achieved through cystectomy, approximately 50% of patients will develop metastases within two years and may ultimately die from the disease. This is likely due to existence of regional or distant microscopic metastatic disease at the time of surgery. (9) The proposed methodology will involve the comprehensive examination of variables associated with patient demographics, tumor characteristics, treatment modalities, and postoperative outcomes. By employing machine learning techniques, we aim to identify key predictors of mortality and post-operative survival, ultimately constructing a model that can inform clinical practice.

## Methods

### Data Collection and Preprocessing

We collected retrospective data on patients with bladder cancer from Fondazione Policlinico Gemelli in Rome, Italy. The dataset included 370 patients with various clinical and pathological variables. Ethical approval was obtained from the institutional review board under protocol number 676-02.

The following variables were considered in the analysis (Table 1)

**Table 1.**
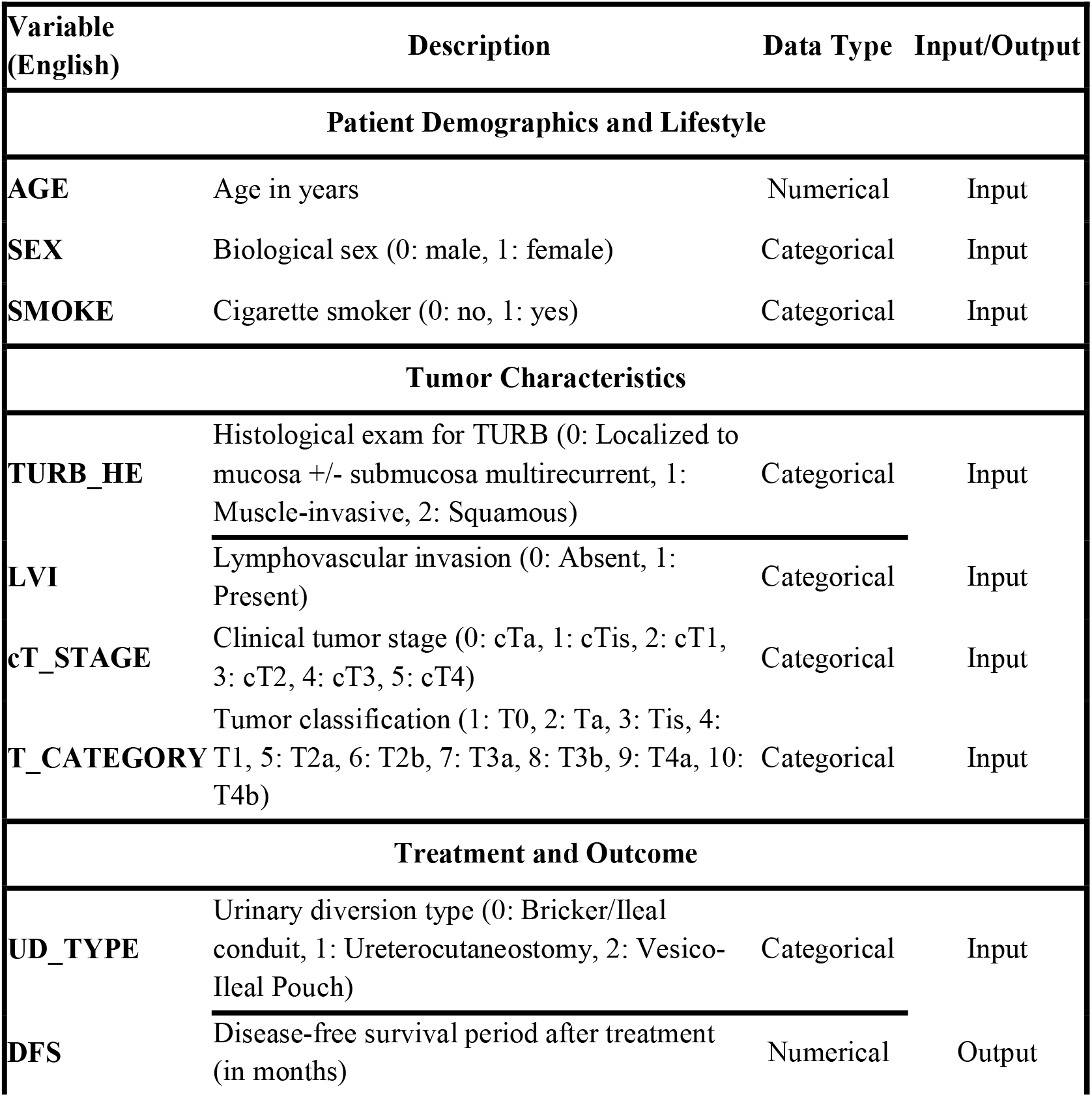
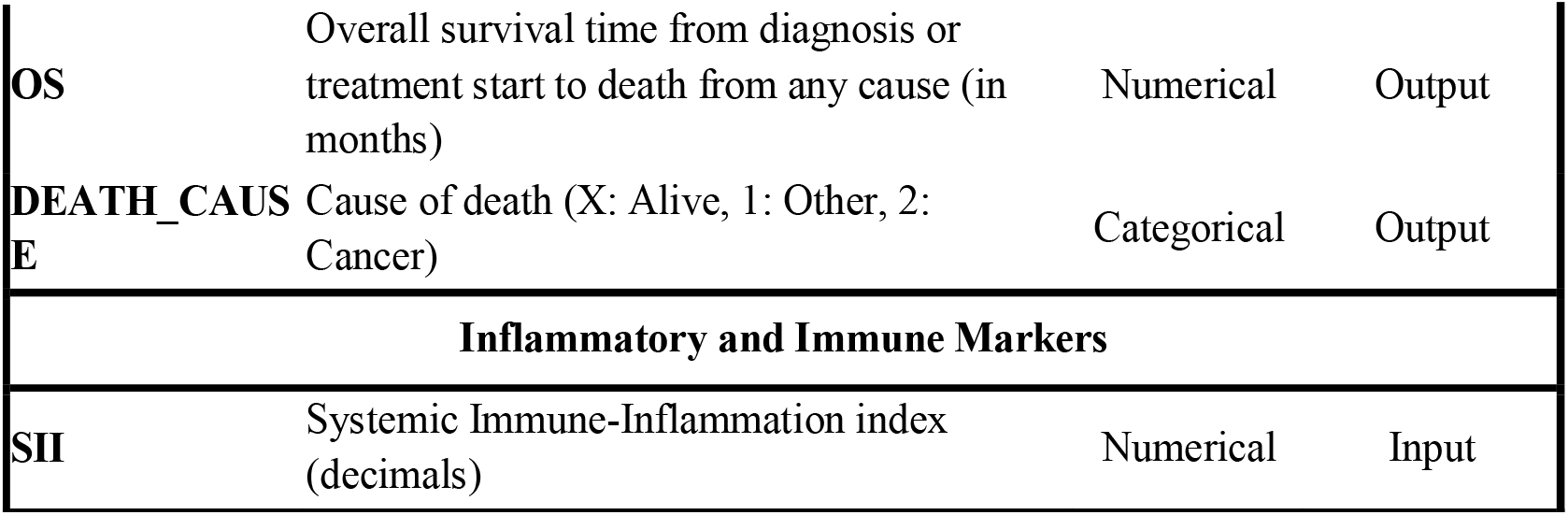
Variables included in the study.

| Variable<br>(English) | Description | Data Type | Input/Output |
| --- | --- | --- | --- |
| Patient Demographics and Lifestyle |  |  |  |
| AGE | Age in years | Numerical | Input |
| SEX | Biological sex (0: male, 1: female) | Categorical | Input |
| SMOKE | Cigarette smoker (0: no, 1: yes) | Categorical | Input |
| Tumor Characteristics |  |  |  |
| TURB_HE | Histological exam for TURB (0: Localized to mucosa +/- submucosa multirecurrent, 1: Muscle-invasive, 2: Squamous) | Categorical | Input |
| LVI | Lymphovascular invasion (0: Absent, 1: Present) | Categorical | Input |
| cT_STAGE | Clinical tumor stage (0: cTa, 1: cTis, 2: cT1, 3: cT2, 4: cT3, 5: cT4) | Categorical | Input |
| T_CATEGORY | Tumor classification (1: T0, 2: Ta, 3: Tis, 4: T1, 5: T2a, 6: T2b, 7: T3a, 8: T3b, 9: T4a, 10: T4b) | Categorical | Input |
| Treatment and Outcome |  |  |  |
| UD_TYPE | Urinary diversion type (0: Bricker/Ileal conduit, 1: Ureterocutaneostomy, 2: Vesico-Ileal Pouch) | Categorical | Input |
| DFS | Disease-free survival period after treatment (in months) | Numerical | Output |
| **OS** | Overall survival time from diagnosis or treatment start to death from any cause (in months) | Numerical | Output |
| **DEATH_CAUSE** | Cause of death (X: Alive, 1: Other, 2: Cancer) | Categorical | Output |
| **Inflammatory and Immune Markers** |  |  |  |
| **SII** | Systemic Immune-Inflammation index (decimals) | Numerical | Input |

The dataset was split into three subsets based on the outcome variables:

1. Disease-free survival (DFS) in months
2. Overall survival (OS) in months
3. Cause of death (alive, other cause, or cancer)

### Data Analysis

We employed multiple machine learning approaches to analyze the relationships between the input variables and outcomes. All analyses were performed using Python with scikit-learn and TensorFlow libraries.

For the regression tasks (DFS and OS prediction):

- Linear regression
- Random forest regression
- Gradient boosting regression
- Support vector regression
- Neural network regression

For the classification task (cause of death prediction):

- Logistic regression
- Random forest classification
- Gradient boosting classification
- Support vector classification
- Neural network classification

### Model Performance Evaluation

For regression tasks, we used mean absolute error (MAE) as the primary metric to compare model performance. For the classification task, we used accuracy and generated confusion matrices to evaluate performance.

### Feature Importance Analysis

For interpretable models like linear and logistic regression, we examined the coefficients to determine the relative importance of input variables. While neural network models are inherently less interpretable, we explored techniques to infer feature importance.

### Cross-validation

To ensure the robustness of our results, we employed k-fold cross-validation for all models.

### Hyperparameter Tuning

For the more complex models (random forests, gradient boosting, and neural networks), we performed hyperparameter optimization using grid search with cross-validation using grid search with cross-validation for the more complex models (random forests, gradient boosting, and neural networks).

### Neural Network Architecture

For the neural network models, we designed a relatively simple architecture given the limited size of our dataset.

### Comparative Analysis

We compared the performance of different models for each task to determine the most suitable approach. Special attention was given to the tradeoff between model performance and interpretability.

## Results

We applied multiple machine learning models to predict three key outcomes: Disease-Free Survival (DFS), Overall Survival (OS), and Cause of Death. The performance of these models was evaluated using appropriate metrics for each task.

### Disease-Free Survival (DFS) Prediction

For DFS prediction, we employed linear regression, random forest, gradient boosting, support vector regression, and a neural network. The performance of these models was evaluated using Mean Absolute Error (MAE).

All models showed similar performance, with MAE values around 22-23 months. The neural network and gradient boosting models slightly outperformed others, but the difference was marginal (approximately 0.4 months). Given the interpretability advantages, we focused on the linear regression model for further analysis.

The linear regression model achieved an MAE of 22.9 months. This indicates that, on average, the model’s predictions deviate from the actual DFS by about 23 months. Considering the distribution of survival times in our dataset, which showed a logarithmic decline with many patients experiencing events in the first few months, this level of accuracy suggests the model captures meaningful patterns in the data.

The coefficients of the linear regression model provided insights into the relative importance of different variables (Figure 1). The analysis of the feature coefficients and intercept provides valuable insights into the linear regression model for Disease-Free Survival (DFS) prediction. The large positive intercept of approximately 30 suggests a baseline DFS prediction, which serves as a starting point before adjusting based on other features. Interestingly, age shows a moderate positive coefficient, indicating that older age is associated with slightly longer DFS, contrary to what might be expected. The clinical T-stage (_cT) emerges as the most influential factor, displaying the most substantial negative coefficient. This implies that higher clinical T-stages are strongly associated with shorter DFS periods.

**Figure 1.**
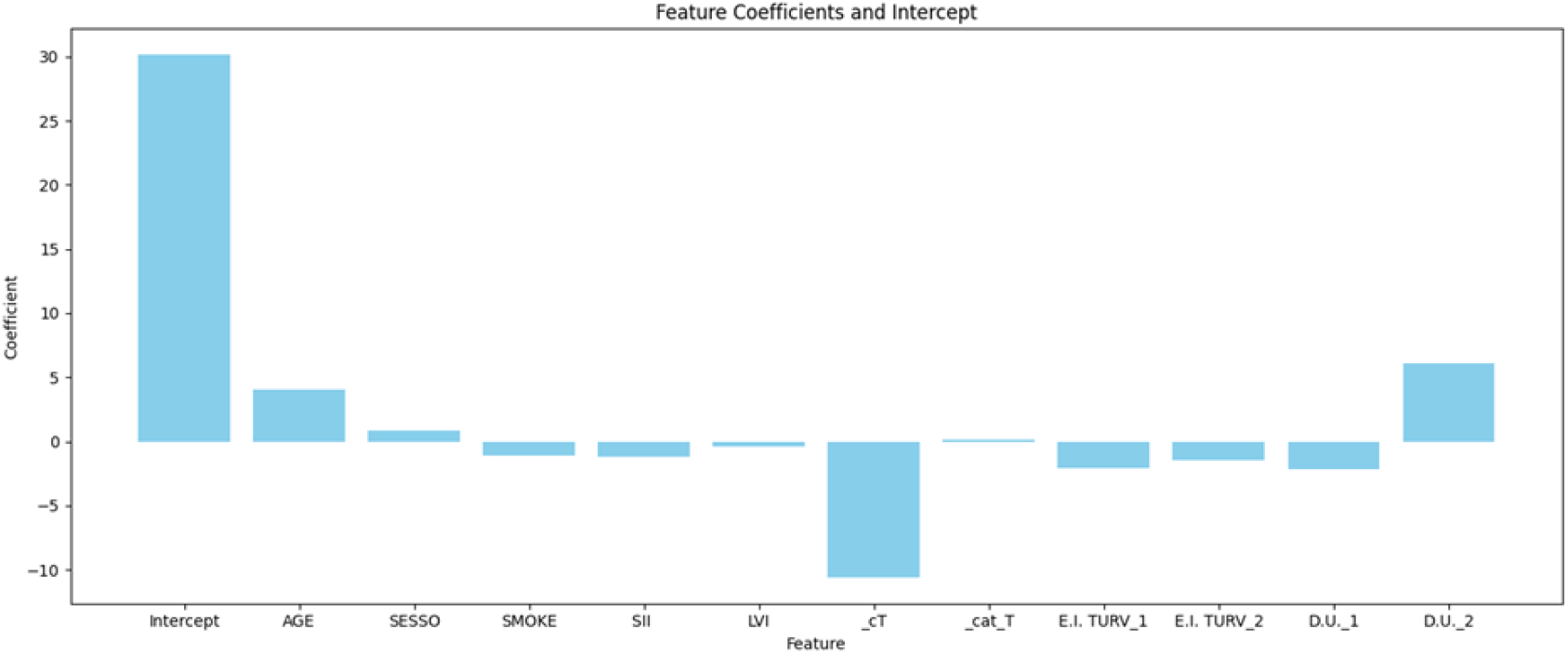
Feature Coefficients and Intercept for Disease-Free Survival (DFS) prediction.

The urinary diversion type, specifically the presence of a vesico-ileal pouch, demonstrates a notable positive coefficient, suggesting that this particular type of urinary diversion may be associated with improved DFS compared to the reference category. The Systemic Immune-Inflammation Index (SII) shows a small negative coefficient, indicating a slight inverse relationship with DFS. Other features such as sex (“SESSO”), smoking status (“SMOKE”), and lymphovascular invasion (“LVI”) exhibit smaller coefficients, suggesting more modest impacts on the prediction model.

### Overall Survival (OS) Prediction

For the overall survival (OS) prediction task, we applied multiple machine learning models, including gradient boosting regression, support vector regression, neural network, random forest regression, and linear regression. The performance of these models was evaluated using Mean Absolute Error (MAE). All models performed similarly, with MAE values around 22-23 months. The gradient-boosting regressor demonstrated the best performance with an MAE of 22.92 months, followed closely by the random forest regressor (MAE = 22.78 months). The neural network (MAE = 23.33 months), support vector regressor (MAE = 23.92 months), and linear regression (MAE = 23.49 months) models showed slightly higher error rates but were still within a similar range.

Different variables showed diverging levels of importance in the linear regression model (Figure 2). The intercept represents the baseline OS prediction when all other variables are at their reference levels. Interestingly, age shows a positive association with OS, suggesting that older patients in this cohort tend to have longer survival. Clinical T-stage emerges as the most influential factor, with a large negative coefficient indicating that higher clinical T-stages are strongly associated with shorter overall survival. The second type of urinary diversion demonstrates a substantial positive coefficient, suggesting it may be linked to improved overall survival compared to the reference category. The Systemic Immune-Inflammation Index (SII) shows a moderate negative association with OS, implying that higher SII values correlate with shorter overall survival. As expected, smoking status exhibits a negative relationship with OS. Other variables, including sex, lymphovascular invasion, and the different categories of E.I. TURV, display smaller coefficients, indicating more modest impacts on the overall survival prediction.

**Figure 2.**
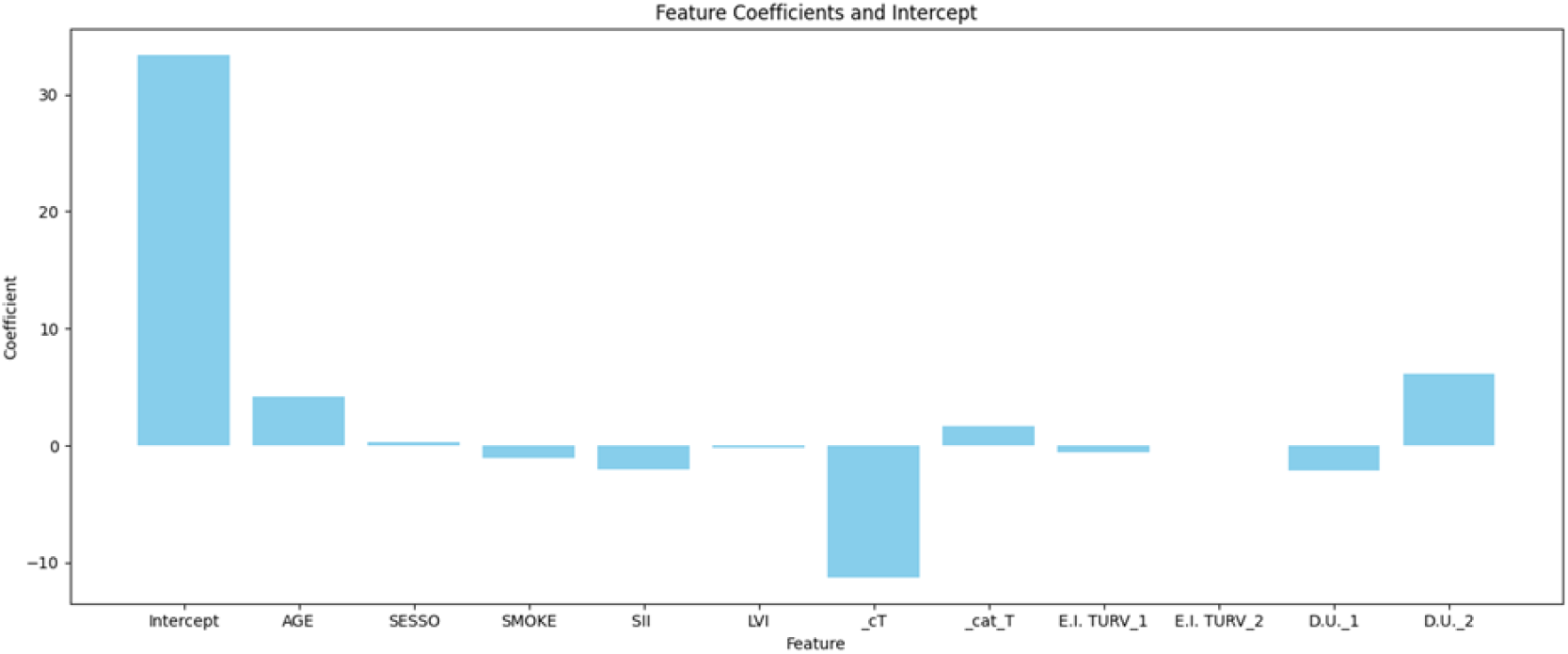
Feature Coefficients and Intercept for Overall Survival (OS) Prediction.

### Cause of Death Classification

To predict the cause of death (cancer-related, other causes, or whether the patient was alive at the time of data collection), we applied multiple machine learning models, including logistic regression, random forest, gradient boosting classifier, support vector classifier, naive Bayes, and a neural network. The performance of these models was evaluated using accuracy scores and confusion matrices (Table 2).

**Table 2.** Model Accuracy Comparison.

| Model | Accuracy | Notes |
| --- | --- | --- |
| **Logistic Regression** | 0.5972 | Not chosen due to consistently predicting class 1, showing poor performance for classes 0 and 2. |
| **Random Forest** | 0.5833 | Not selected as it consistently predicted class 0, failing to distinguish between different causes of death. |
| **Gradient Boosting Classifier** | 0.5000 | Rejected due to always predicting class 0, showing no ability to differentiate between outcomes. |
| **Support Vector Classifier** | 0.6667 | While accuracy matched the neural network, it never predicted class 1, limiting its usefulness. |
| **Naive Bayes** | 0.6111 | Rejected due to always predicting class 0, showing no ability to differentiate between outcomes. |
| **Neural Network** | 0.6667 | Chosen as the best model due to its balanced performance across classes, especially in distinguishing between alive patients (class 0) and cancer deaths (class 2). |

The neural network model demonstrated the best overall performance for this task, achieving an accuracy of 0.6667. This means that the model correctly predicted the cause of death for approximately 66.67% of the cases in the test set. While this accuracy is moderate, it represents a significant improvement over random guessing (which would yield an accuracy of about 33% for a three-class problem).

Table 3 highlights the comparative performance of machine learning models across the three prediction tasks: Disease-Free Survival (DFS), Overall Survival (OS), and Cause of Death Classification, emphasizing the tradeoff between performance and interpretability in clinical applications. While the Neural Network and Gradient Boosting models demonstrated the lowest Mean Absolute Error (MAE) for DFS and OS predictions, the Neural Network emerged as the most balanced and accurate model for cause-of-death classification with an accuracy of 66.67%. Linear Regression, despite slightly higher MAE values, was highlighted for its interpretability, particularly in analyzing feature importance.

**Table 3.**
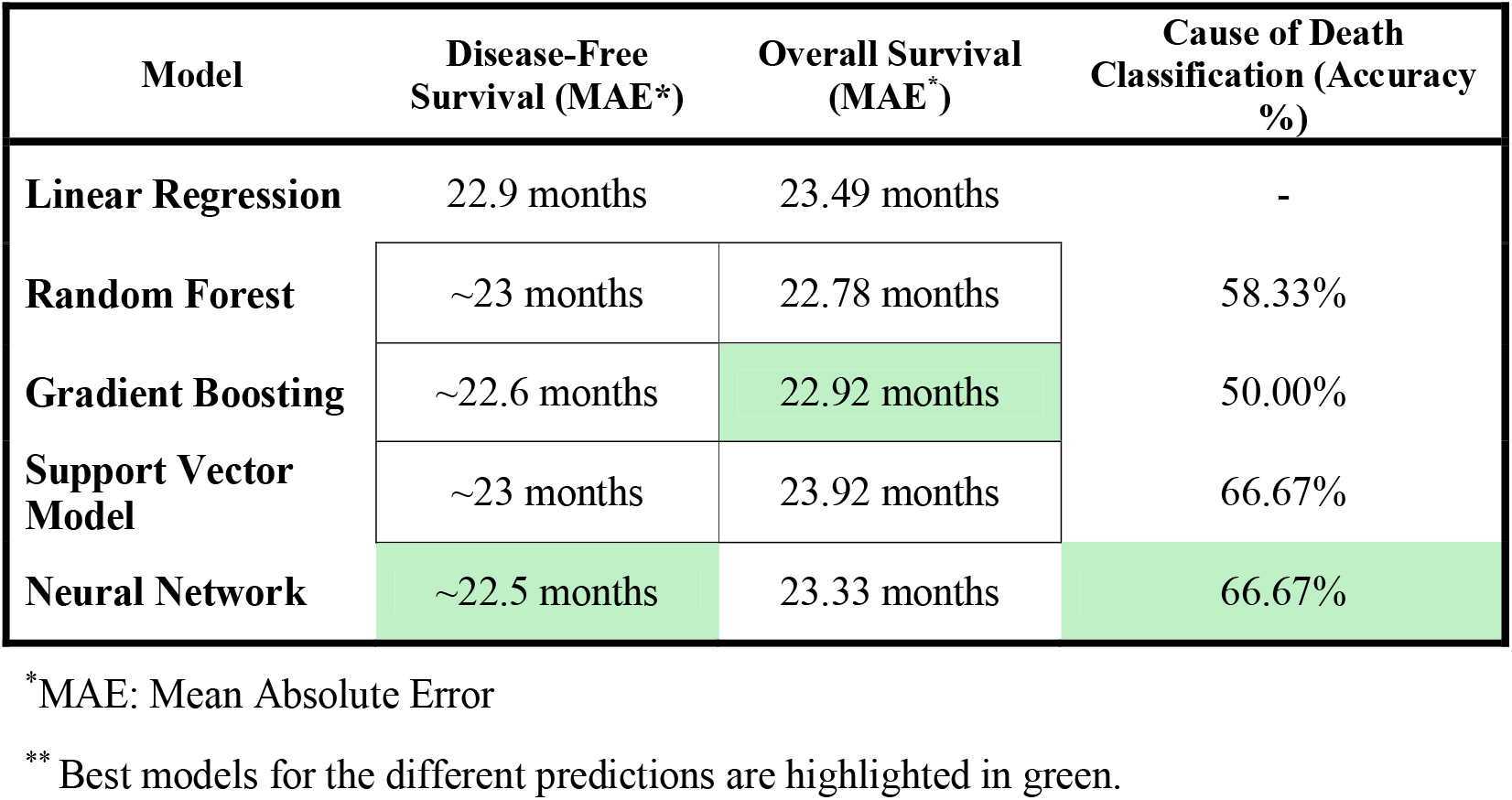
Model Performance Comparison Across Prediction Tasks.

| Model | Disease-Free Survival (MAE*) | Overall Survival (MAE*) | Cause of Death Classification (Accuracy %) |
| --- | --- | --- | --- |
| **Linear Regression** | 22.9 months | 23.49 months | - |
| **Random Forest** | ~23 months | 22.78 months | 58.33% |
| **Gradient Boosting** | ~22.6 months | 22.92 months | 50.00% |
| **Support Vector Model** | ~23 months | 23.92 months | 66.67% |
| **Neural Network** | ~22.5 months | 23.33 months | 66.67% |

\* MAE: Mean Absolute Error
\*\* Best models for the different predictions are highlighted in green.

The confusion matrix heatmap for the neural network model visually represents these results, with darker blue colors indicating higher prediction accuracy for each class (Figure 3). It revealed high accuracy in predicting patients still alive (class 0) and cancer-related deaths (class 2), while it has poor performance in predicting deaths from other causes (class 1). This suggests that the model is particularly effective at distinguishing between patients who are still alive and those who died from cancer but struggles to accurately identify deaths from other causes.

**Figure 3.**
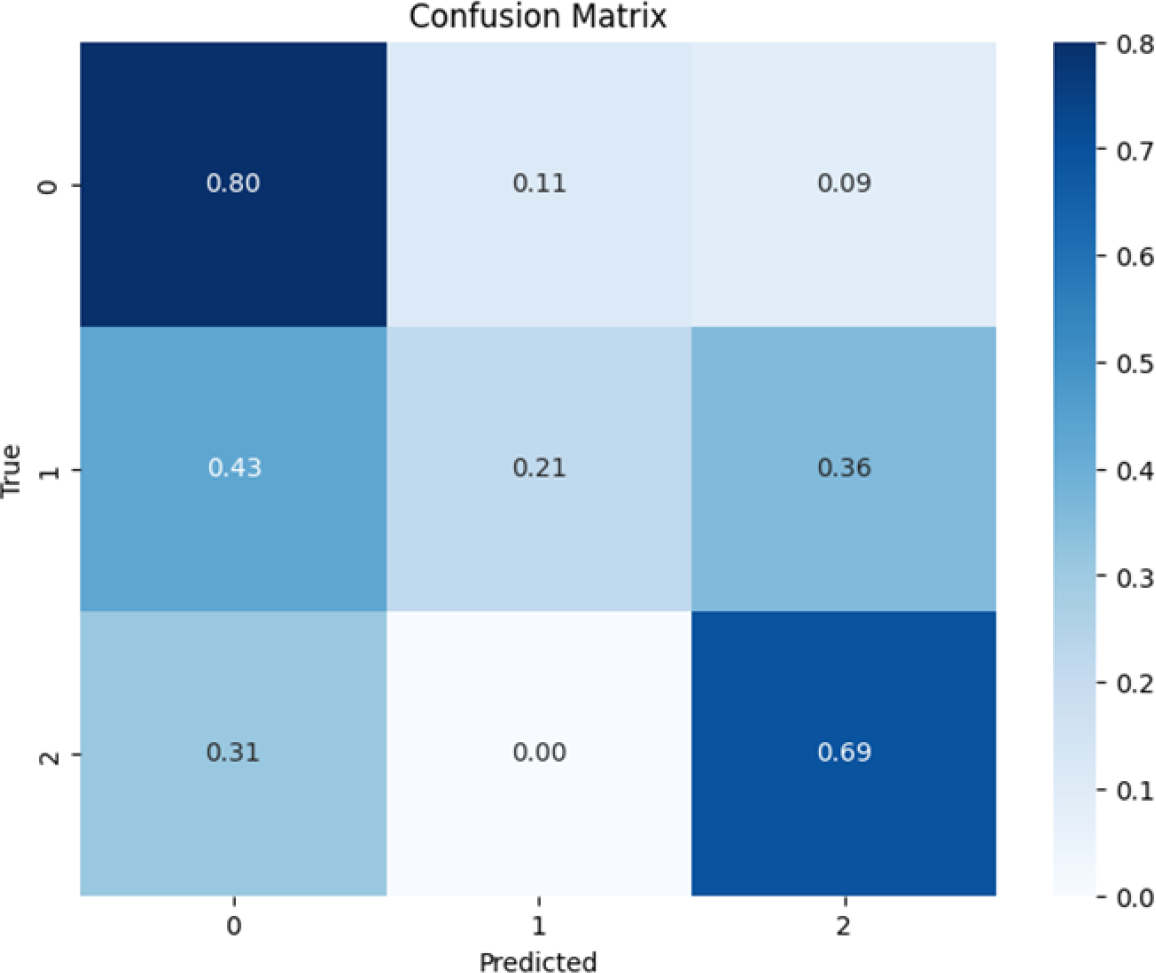
Confusion Matrix for the Neural Network Model

## Discussion

This study presents the development of a survival prediction model in bladder cancer patients using machine learning approaches. Our findings demonstrate that modern predictive algorithms show promising accuracy in forecasting disease-free survival, overall survival, and cause of death. The limited sample and the paucity of included categories for the analysis suggest predictive algorithms trained with additional resources might significantly improve in the demonstrated accuracy.

The observation that age showed a positive correlation with survival outcomes is particularly intriguing and seemingly counterintuitive. This “age paradox” might be explained by several factors. First, older patients often receive more conservative treatment approaches, potentially leading to selection bias in surgical candidates. Additionally, younger patients with bladder cancer often present with more aggressive disease variants, which could account for their relatively poorer outcomes. (10)

The emergence of clinical T-stage as the strongest predictor aligns with established prognostic factors in bladder cancer. (11) Additionally, inflammatory markers, particularly the Systemic Immune-Inflammation Index (SII), show a negative correlation with survival outcomes and support recent findings in other malignancies. (12) This relationship likely reflects the complex interplay between systemic inflammation and cancer progression, where elevated SII indicates a pro-tumoral inflammatory state. (13)

The impact of urinary diversion type on survival outcomes represents an interesting finding. Previous studies had found a protective effect of orthotopic neobladder reconstruction against urethral recurrence in male patients undergoing radical cystectomy for bladder cancer. While that was not present in our patients, we found a positive association between vesico-ileal pouch construction and improved survival. This association may reflect both patient selection and the physiological advantages of this approach; however, this observation should be interpreted cautiously, as confounding factors such as surgical expertise and patient characteristics that were not considered in the present study could influence these results.

Our machine learning models achieved prediction accuracies comparable to previous studies. The neural network’s 66.67% accuracy in cause-of-death prediction, while modest, represents an encouraging amount, given the restricted resources and the paucity of categories considered for the analysis, when compared with studies published a few years ago with samples that were significantly higher and marginally higher accuracy in mortality and recurrence prediction. (14) A recently published systematic review investigating machine learning algorithms for bladder cancer cystectomy outcomes found that most of the algorithms would not exceed 70% accuracy, sometimes even 60%. (15) The integration of SII into predictive models represents a particularly promising direction. As a low-cost, readily available biomarker, SII could enhance current prognostic tools without adding significant complexity or cost to patient evaluation. (16)

A notable limitation of our study is the relatively high MAE values (22-23 months) in survival predictions. These MAE values make the algorithm unsuitable for precise individual patient counseling or treatment planning where accurate timing is critical, such as patients in emergency settings or showing postoperative complications. (17) However, this level of accuracy remains acceptable for clinical trial patient stratification and allocation, particularly in trials where broad risk categories rather than precise survival estimates are needed for randomization, such as balancing treatment arms in clinical trials by identifying comparable risk groups or supporting enrollment decisions in competing risks studies where precise timing is less critical than overall risk assessment. (18)

### Limitations and reproducibility

While we don’t have direct access to feature coefficients for the neural network model, the performance across different models suggests that the input variables contain meaningful predictive information for distinguishing between alive patients and those who died from cancer. However, the consistent difficulty in predicting deaths from other causes across all models indicates that our current set of predictors may not adequately capture the factors leading to non-cancer deaths in this patient population.

#### Model Comparison and Robustness

Across all three prediction tasks, we observed consistent performance among different model types. This consistency suggests that we have likely reached the predictive limit of the current dataset. Similar performance across various algorithms, from simple linear models to complex neural networks, indicates that the predictive signal in the data has been effectively captured.

#### Code and Reproducibility

All code used for this analysis is available at https://tinyurl.com/486yzuy4 to ensure the reproducibility of our results.

## Conclusion

Our study demonstrates the potential utility of machine learning approaches in predicting bladder cancer outcomes following cystectomy. While the achieved accuracy levels are modest, with MAE values of 22-23 months for survival predictions and 66.67% accuracy in cause-of-death classification, they align with current literature benchmarks and provide a foundation for future development. The identification of clinical T-stage as the primary predictor and the consistent negative correlation of SII with survival outcomes validate these parameters as valuable prognostic indicators. The current model’s performance, though not suitable for precise individual prognostication, shows particular promise for clinical trial stratification and cohort allocation. Future studies with larger datasets and additional predictive variables may enhance the model’s accuracy and broaden its clinical applications. Integrating readily available biomarkers like SII represents a cost-effective approach to improving prognostic tools. These findings contribute to the growing body of evidence supporting the role of machine learning in oncological decision-making while acknowledging the need for continued refinement and validation in larger cohorts.

## Authors contributions

FAC, BR, and PR elaborated on the first manuscript concept. AN and FAC performed the statistical analysis. FAC, VDV and PR wrote the article. MS, NF, and GM, reviewed and approved the final manuscript.

